# The attitudes, perceptions and experiences of medical school applicants following the closure of schools and cancellation of public examinations in 2020 due to the COVID-19 pandemic

**DOI:** 10.1101/2020.06.02.20116855

**Authors:** Katherine Woolf, Dave Harrison, I C McManus

## Abstract

**Objective:** To describe medical applicants’ experiences of education and their views on changes to medical school admissions, including the awarding of calculated grades, following the 2020 closure of schools and universities, and the cancellation of public examinations in the United Kingdom due to the COVID-19/coronavirus pandemic. To understand how applicants from diverse social backgrounds might differ in these regards.

**Design:** Cross-sectional questionnaire study forming part of the longitudinal United Kingdom Medical Applicant Cohort Study (UKMACS).

**Setting:** United Kingdom medical school admissions.

**Participants:** 2887 participants (68% female; 64% with at least one degree-educated parent; 63% with at least one parent in the highest socioeconomic group) completed an online questionnaire between 8^th^ and 22^nd^ April 2020. To be invited to complete the questionnaire, participants had to have registered to take the University Clinical Admissions Test (UCAT) in 2019 and to have agreed to be invited to take part in the study, or they needed to have completed one or more previous UKMACS questionnaires. They also need to have been seriously considering applying to study medicine in the UK for entry in 2020 between May and October 2019, and be resident in the UK or Islands/Crown Dependencies.

**Main outcome measures:** Views on calculated grades, views on potential changes to medical school admissions and teaching in 2020 and 2021, reported experiences of education following the closure of educational institutions in March 2020.

**Results:** Respondents had concerns about the calculated grades that will replace A-level examinations, especially female applicants and applicants from Black Asian and Minority Ethnic (BAME) backgrounds who felt teachers would find it difficult to grade and rank students accurately, as well as those from non-selective state schools and those living in deprived areas who had some concerns about the grade standardisation process. Calculated grades were not considered fair enough by a majority to use in the acceptance or rejection of medical offer-holders, but several measures - including interview and aptitude test scores - were considered fair enough to use in combination. Respondents from non-selective state (public) schools reported less use of and less access to educational resources compared to their counterparts at private/selective schools. In particular they reported less online teaching in real time, and reported spending less time studying during the lockdown.

**Conclusions:** The coronavirus pandemic will have significant and long term impacts on the selection, education and performance of our future medical workforce. It is important that the views and experiences of medical applicants from diverse backgrounds are taken into consideration in decisions affecting their futures and the future of the profession.

## Introduction

The UK Medical Applicant Cohort Study (UKMACS) is a study of United Kingdom (UK) medical school admissions. It is primarily a longitudinal questionnaire study of UK residents who in the summer and autumn of 2019 were seriously considering applying to study medicine in the UK for entry in 2020. UKMACS questionnaire data are subsequently linked to administrative data on all UK medical applicants held within the UK Medical Education Database (www.ukmed.ac.uk). Wave 1 data were collected between May and September 2019 and asked how applicants from different backgrounds were choosing which medical schools to apply to. Wave 2 data were collected from November 2019 to January 2020 and asked which medical schools and universities participants had applied to and how they had made their choices.

In March 2020 it was announced that UK schools would close and A-level (and equivalent public examinations) would be cancelled due to the coronavirus/COVID-19 outbreak in the UK. This was one of the most major disruptions ever to affect education and university admissions in the UK and was very significant for the UKMACS cohort, who are mostly in their final year of schooling and were due to sit examinations in the summer of 2020.

We therefore decided to administer an additional unplanned UKMACS questionnaire to understand what medical applicants were experiencing in terms of education, their views on how grades would be awarded following cancellations of public examinations, and their views on how medical schools might respond with regard to admissions policies. We particularly sought to understand how applicants from diverse social backgrounds might differ in these regards with the aim of facilitating the inclusion of applicant perspectives and experiences in discussions about changes to medical school admissions and medical education.(1)

### Calculated grades

The absence of A-levels and other equivalent public examinations in March 2020 meant that alternative methods of assessment for candidates had to be found, not least as A-levels are “the single most important bit of information [used in selection]” by universities.(2) On April 3^rd^ *Ofqual* (Office of Qualifications and Examinations Regulation) in England announced that A-level, GCSE and other exams under its purview in England would be replaced by calculated grades based on teachers estimation of the grades that their students would have attained, which would then be standardised centrally.(3) The Scottish Qualification Authority (SQA) and other national bodies also announced similar processes for their examinations.

Performance in A-level examinations has long-term impacts (4, 5) making changes to how grades are awarded potentially very significant. The use of calculated grades in place of exam grades raises many questions, some of which were summarised in a letter to *The Guardian* by Yasmin Hussein, a GCSE student in Birmingham, who said that,

> “… the … exam hall [is] a level playing field for all abilities, races and genders to get the grades they truly worked hard for and in true anonymity (as the examiners marking don’t know you). [… Now we] are being given grades based on mere predictions.” Yasmin Hussein, letter to The Guardian, March 29th 2020.(6)

Among teachers, survey data suggests that there are doubts about the accuracy and fairness of calculated grades, with 39% saying that all students would get a fair deal, 24% saying they would not, and 37% not knowing or not answering. There were also doubts about fairness for students from Black Asian and Minority Ethnic (BAME) backgrounds, about those working hard in the last weeks before an exam being penalised, about teacher ‘favouritism’, although there were others who commented that the process is as fair as possible under the circumstances.(7)

Concerns are also present amongst applicants to universities. In a survey carried out by HEPI (Higher Education Policy Institute) before the details of calculated grades were announced, but after it was known that grades would in some way be predicted, 27% thought that their predicted grades were worse than they were likely actually to have attained, compared with 13% thinking their predicted grades were better than they would actually attain.(8)

Another survey of 511 university applicants (including 452 A-level students) conducted for the Sutton Trust found that just under half believed the new A-level grading system would result in their receiving poorer grades but working class respondents were more worried about large negative consequences compared to middle class students. Nearly three quarters believed the new system was less fair than examination grades and interestingly this was more of a concern for applicants from higher socioeconomic backgrounds. Nearly half of applicants felt the COVID-19 crisis would negatively impact their chances of getting into their first choice university, and this was more common among working class respondents.(9)

The impact on medical school admissions of examination cancellations and their replacement with calculated grades is, at the time of writing, still not completely clear. *Ofqual* states that,

> “The grades awarded to students will have equal status to the grades awarded in other years and should be treated in this way by universities, colleges and employers. On the results slips and certificates, grades will be reported in the same way as in previous years”.(3), p.6.

The decisions of *Ofqual* in this case are in effect governmental decrees, supported by Ministerial statement, and universities and other bodies will therefore abide by them, as was affirmed by the Medical Schools Council on 5^th^ May 2020.(10) That does not mean however that other factors may not need to be taken into account in some cases, as for instance when applicants do not attain the grades needed for their conditional offers, or for applicants in clearing. Furthermore in guidance updated on 1^st^ May 2020 the Government stated that “if a student does not feel their grade reflects their performance, they will have the opportunity to take an exam in the autumn”(11) with *Ofqual* expanding on 15^th^ May 2020 that “students will be able to use the higher of the two grades for future progression.”(3) This raises questions for university admissions, as Medical Schools Council acknowledged in their statement of 5^th^ May 2020:

> “There are a number of issues that the education sector as a whole is yet to resolve. These include how appeals against calculated grades will work across the UK and when students will be able to sit exams if they are unhappy with their calculated grade. The impact of these issues on medical admissions is unclear but medical schools are actively engaging in these discussions and are working hard to develop solutions that are fair to applicants.”(10)

### Education during the pandemic

As well as examinations being cancelled, UK schools closed on 20^th^ March 2020 to all except the children of critical workers and vulnerable children. While primary schools in England reopened to some year groups on 1^st^ June 2020, this is not the case in other UK countries and in practice it seems likely that secondary schools and colleges will remain closed to most students until September 2020. Similarly in mid-March 2020 many universities suspended face-to-face teaching for the academic year 2019/2020, and on 19^th^ May 2020 Cambridge University announced that all lectures would be online until academic year 2021/2022 with many others likely to follow suit.

The impact of school closures on student learning and outcomes will be significant (12-14) and there is emerging evidence that it may be particularly problematic for those from poorer backgrounds and/or at state-funded schools. The Institute of Fiscal Studies analysed survey data from a weighted sample of over 4000 parents with children aged between four and 15 years old (15). Parents were asked about the types of home learning their child was doing, the resources provided by the school, and resources available at home for learning in the first two weeks of May 2020. Among secondary school children, those from the richest quintile were spending on average slightly over an hour more per day on learning compared to those in the poorest quintile, amounting to several weeks more learning over the course of the time schools are likely to be closed. Indeed, children in the richest families were spending significantly more time than others on *all* educational activities, but particularly those provided by schools and from private tutors. Even among state school pupils, children from the richest families were reported to have greater access to face-to-face online teaching, which the report authors argue is likely to be of higher educational value than other resources that require more parent input, particularly since the poorest parents of secondary school children were less likely to find it easy to support their child’s home learning.

The results of the IFS report chime with data from *Teacher Tapp*, an ongoing weighted survey of several thousand teachers in England.(16) At the start of the lockdown (23^rd^ March 2020) private school secondary schools were much more likely than state secondary schools to be using online videoconferencing (27% vs 2%) and online chat (18% vs 3%). The above-mentioned Sutton Trust report (9) also found socioeconomic differences in access to “internet access, devices for learning or a suitable place to study” and differences in the amount of A-level teaching being conducted by teachers at private and state schools.

Among those secondary school pupils who had applied to university, the Sutton Trust report authors argued that students from lower socio-economic backgrounds are also likely face additional disadvantages both with their university applications and when starting university:

> “Given the uncertainty caused by these changes [to education resulting from COVID-19], university applicants are likely to need more support than ever to navigate the process [of applying to university]. This will be even more important for young people from lower socio-economic backgrounds, who are less likely to be able to draw on the advice of family members with higher education experience themselves. But with schools closed for most pupils, it may be difficult for applicants to get the help they need. Similarly, there’s also a danger that this year’s applicants will miss out on A level content during the lockdown […]. For disadvantaged students about to go on to higher education, this could leave them with gaps in their knowledge base, putting them behind their peers before they have even begun at university.” [p1. (9)]

### The present study

This study aimed to explore and describe perceptions of calculated grades, of student selection more generally, and of educational experiences during school and university closures, in a large group of medical school applicants, who are typically high-attaining students. A range of background factors were assessed to determine how perceptions differed according to demographic and other measures. Data collection took place between April 8^th^ and April 22^nd^, which was about two and a half weeks after school closures.

## Methods

### Study design

Cross-sectional questionnaire study, which formed part of the longitudinal UK Medical Applicant Cohort Study.

### Eligibility

To be invited to complete the questionnaire, participants had to have registered to take the University Clinical Admissions Test (UCAT) in 2019 and to have agreed to be invited to take part in UKMACS, or they needed to have completed one or more previous UKMACS questionnaires. They also need to have been seriously considering applying to study medicine in the UK for entry in 2020, and be resident in the UK or Islands/Crown Dependencies.

The following groups were excluded from the study and not sent an invite:

- those who had previously requested their data be removed from the UKMACS database;
- previous UKMACS respondents who had not agreed to be contacted for further research;
- previous UKMACS respondents who had previously not consented to having their personal information retained by the research team;
- previous UKMACS respondents who had previously not consented to their personal information being linked with other information for research purposes.

### Questionnaire development

During the development of the questionnaire *Ofqual* announced that calculated grades would be awarded. We were therefore able to assess perceptions of how calculated grades would be awarded and used, and perceptions of other possible methods medical schools could use to select or reject offer-holders. We also about potential knock-on effects of calculated grades, including what medical schools should do if they have more applicants meeting their offers than they have medical school places, and how rejected applicants should be treated in the 2021 application cycle.

With uncertainty about whether medical schools and universities would be able to open at the usual time in academic year 2020/2021 we asked applicants whether medical schools should defer opening until teaching could be done face-to-face, or whether they should open online.

We asked applicants about their use of educational resources provided by schools/colleges/universities, what preparation they were doing for university/medical school, and how much time they were spending on various activities including studying, caring, and volunteering (clinical and non-clinical).

We included self-reported measures of academic attainment and socio-demographic measures we had used in previous UKMACS questionnaires, as well as the 15-item Big Five personality measure used in the national longitudinal cohort study *Understanding Society* [https://www.understandingsociety.ac.uk/documentation/mainstage/dataset-documentation/term/personality-traits].

Most questions were designed specifically for this questionnaire since they asked about unprecedented events and validated items were not available. We constructed the questionnaire with JISC Online Surveys [https://www.onlinesurveys.ac.uk/] and piloted the questionnaire and information sheet with two current applicants to medical school. Amendments were made in response to feedback from the applicants and from Medical Schools Council.

### Questionnaire administration

Participants were sent an email invitation and link to the current questionnaire on the afternoon of 8^th^ April 2020. 18,665 invitations were sent, with up to two email reminders and two text message reminders. The questionnaire closing date was 20^th^ April 2020, with responses accepted up to 22^nd^ April 2020.

The questionnaire was administered on the JISC Online Surveys platform. All participants were given the option to withdraw from the study and to request that their contact data be removed from the participant list. Any participant who had responded but then wished for their responses to be removed from the study were able to do this by contacting the UKMACS Research Team by the end of 22^nd^ April 2020. No participants requested that their questionnaire responses be removed.

### Statistical analysis

Descriptive and univariate analyses were performed in SPSS v26. Multivariate analyses were performed in R, with missing values handled using multiple imputation using the *mice* package.(17) Following the general advice of van Buuren (18) missing values were calculated using *pmm* (predictive mean matching), which as van Buuren says, is a good “ all-round method with exceptional properties” (p.84). *pmm* is the default method in the *mice()* function for all scale types (binary, ordinal, numeric) and has the advantage that imputed values are always taken from the existing range of actual values in the data, with *pmm* being robust against mis-specification (pp77- 84). The number for the pool of candidate donors, d, was set at 5, the default in *mice()*, and the number of imputations, m, was set at 25. Regression analyses on the 25 *mira* datasets were carried out using the *lm()* function within the *with()* function, and separate sets of results in the *mipo* dataset were combined with the *pool()* function.

Multiple regression analysis entered all socio-demographic and educational predictor variables into the analysis simultaneously, and results are only reported which were significant with p<.01 after taking all other variables into account, so the analysis is relatively conservative. The nine socio-demographic and educational variables used were: ethnicity, gender, school type, parental higher education, IMD quintile, mean GCSE points, mean top three predicted A-levels, UCAT score, number of medical school offers.

### Ethics and data protection

The study was approved by the UCL Research Ethics Committee Chair on 8^th^ April 2020 as an amendment to the ongoing UKMACS longitudinal questionnaire study (reference: 0511/014).

### Funding

Katherine Woolf and David Harrison are funded by a National Institute for Health Research (NIHR) Career Development Fellowship for this research project. This publication presents independent research funded by the National Institute for Health Research (NIHR). The views expressed are those of the author(s) and not necessarily those of the NHS, the NIHR or the Department of Health and Social Care.

## Results

### Participants

3071 participants completed the questionnaire, of whom 2904 stated they were eligible to take part (i.e. seriously considering applying to study medicine in the UK in 2020 and resident in the UK or Islands/Crown Dependencies). After removing 16 respondents who did not consent to have their data analysed and 11 duplicates, there were 2877 valid cases for analysis, which is 15% of those invited. This is subsequently referred to as the full sample.

The main analyses were performed on a restricted sample of 1562 respondents currently in Year 13, who had applied to medicine for entry in 2020, with at least three predicted A-levels and no achieved A-levels. Results are also reported in the Supplementary files for respondent groups excluded from the restricted sample, notably those living in Scotland and those not currently in Year 13 - see Supplementary files 1 and 2.

### Missing data

The analysis considered 120 measures in the restricted sample. The data could be divided into three groups:

1. *Questionnaire items*. The questionnaire asked about attitudes to 98 different topics concerning medical school entrance. Of 153,076 data points, 10788 (7.2%) were missing. For the individual variables, the median percentage of missing data values was 0.48%, with 75 measures having fewer than 5% of missing values.
2. *Derived measures*. Factor scores were calculated for the 98 attitudinal questionnaire items, using PFA in the SPSS Factor program, followed by Varimax rotation. A scree-slope analysis suggested that six factors were present, and these were extracted using the regression method. Given the small proportion of missing values, mean substitution was used for missing values, with the outcome that none of the nine derived factor scores had missing values.
3. *Demographic and educational items*. For 12 demographic measures, 462 of 18744 measures were missing (2.5%), with a median of 1.0% per measure, and 11 measures having fewer than 5% missing values. Ethnic origin was not asked about in the present study. The ethnicity of 889 respondents who had reported their ethnicity in a previous questionnaire were imported into the present dataset; 43.1% of ethnicity measures were therefore missing. IMD_Quintile was obtained from postcodes in England, Wales and Scotland, and was missing in 14.8% of cases.

There were four educational attainment items (top three predicted A-level grades, UCAT score, BMAT score, and mean GCSE grade). Top three predicted A-level grades were present for all because the sample was based on that criterion. Of the remaining three measures, 1852 out of 4686 (39.5%) were missing: UCAT scores were missing in 13.6% of cases, and BMAT scores in 61.3% of cases, but in both cases missing values were mostly structurally missing, candidates mostly having taken only one aptitude test or the other. Mean GCSE grade was missing in 43.1% of cases, having been imported from a previous UKMACS questionnaire.

Participants self-reported their current or most recent school in the current questionnaire. This question was also present in the Wave 1 UKMACS questionnaire. For schools in England, publicly-available administrative data were available on school type (e.g. independent, voluntary aided) and for state-funded schools there were data on whether the admissions policy was selective or non-selective. These were combined to create a binary variable of School Type (non-selective state schools vs private/selective schools) for 1132 respondents (27.1% missing). A composite variable was created using present responses and the responses in the Wave 1 questionnaire, so that data were available for 1158 respondents with values missing in 25.9% of respondents.

### Demographics

Demographics for the full and restricted samples are reported in **Figure 1**.

**Figure 1.**
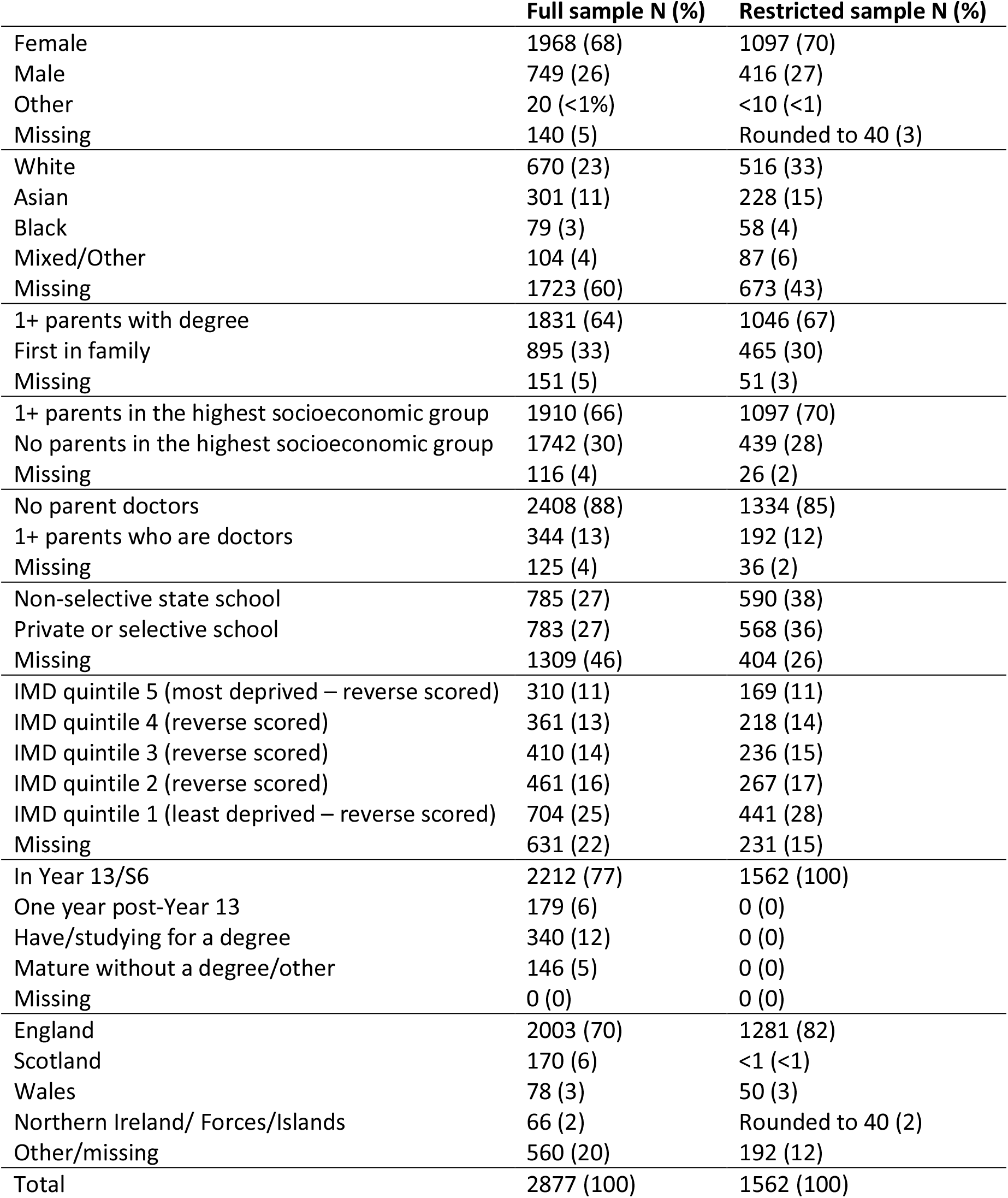
***Demographics for the full sample and the restricted sample (of those in Year 13, with at least three predicted*** A-level***s, no achieved*** A-level***s, who had applied to study medicine). Rounding to prevent identifying individuals***.

### Education and achievement

#### Predicted A-levels

A-level grades were scored as A*=12, A=10, B=8 etc. Predicted A-levels were sometimes reported as being between two grades e.g. A*/A was scored 11, A/B was scored 9, etc. The mean predicted A-level grades were calculated for the top three grades regardless of subject *(Mean top three predicted* A-levels), and for all grades *(Mean predicted* A-levels). *Mean top three predicted* A-levels was 10.89 and *Mean predicted* A-levels was 10.71, both of which are over an A grade.

#### UCAT, BMAT, GAMSAT

1546 participants (99.1%) reported having taken UCAT; 765 (49.0%) reported having taken BMAT; and none reported having taken GAMSAT. Of the 1350 participants who reported a total UCAT score that was greater than 1799 and less than 3601, the mean score was 2660 (SD=235).

#### GCSE

GCSE grades can range from 1 to 9. A variable *Mean GCSE* was calculated by dividing the total GCSE points by the number of GCSEs taken, and the mean was 7.91 (SD=-.71).

#### Relationships between educational measures

UCAT score correlated with *Mean top three predicted* A-levels at 0.418 (p<.001) and with *Mean GCSE* at 0.487 (p<.001). *Mean GCSE* and *Mean top three predicted* A-levels correlated at 0.611 (p<.001).

Participants at non-selective state schools had lower scores on all attainment measures *(Mean GCSE:* difference=0.3 points, p<.001; *Mean top three predicted* A-levels: difference=0.23 points, p<.001; UCAT score: difference=89 points, p<.001).

### Medical school offers

1292 (85%) respondents had applied to four medical courses, 1289 (82.5%) had at least one offer, 177 (11.3%) had four offers, and 204 (13%) were waiting to hear from at least one medical school at the time of completing the questionnaire.

Respondents who did not have a parent/carer with a university degree were less likely to have a medical offer (78.1% vs 85.0%; p=0.001).

### Applicant views on admissions

#### Perceptions of the fairness of methods medical schools could consider using in the selection of offer-holders

Participants were asked to rate the fairness of 17 measures, including calculated grades, that medical schools could potentially use to decide to accept or reject offer-holder following exam cancellations. Rating categories were: “Unfair: should not be used” “Quite unfair: avoid if possible” “Quite fair: could be used in combination with other measures” “Very fair: could be used alone”. Participants could also type comments and suggestions in a freetext box.

No single measure was felt by a majority of participants to be fair enough to use on its own. The method rated by the most participants as fair enough to use alone was *Exam grades taken in September 2020 (if these take place)* which 32.3% rated as very fair. This was followed by *Predicted Grades declared on UCAS application* (26.2% very fair), *Calculated grades* (22.6% very fair), *GCSE grades* (20.4% very fair) and *Score at interview* (19.5% very fair).^1^

By contrast, several methods were felt by a majority of participants to be fair enough to be used in combination with other methods. Three methods were felt by three quarters of respondents to be quite or very fair: *Predicted grades* (80.6%), *GCSE grades* (73.8%), and *Score at interview* (73.4%). A further four methods were felt by two thirds of respondents to be quite or very fair: *Calculated grades* (68.8%), *UCAS teacher reference* (63.5%), *UCAS personal statement* (62.5%) and *BMAT score* (62.5%). A further three methods were felt by over half of respondents to be quite or very fair: *UCAT score* (57.1%), *Exams taken in September 2020* (56.7%), and AS *levels taken in Year 12* (52.7%). At the other end of the scale, only a fifth (20.3%) of participants felt *Attendance at widening participation activities* was quite fair or very fair. See **Figure 2**.

**Figure 2:**
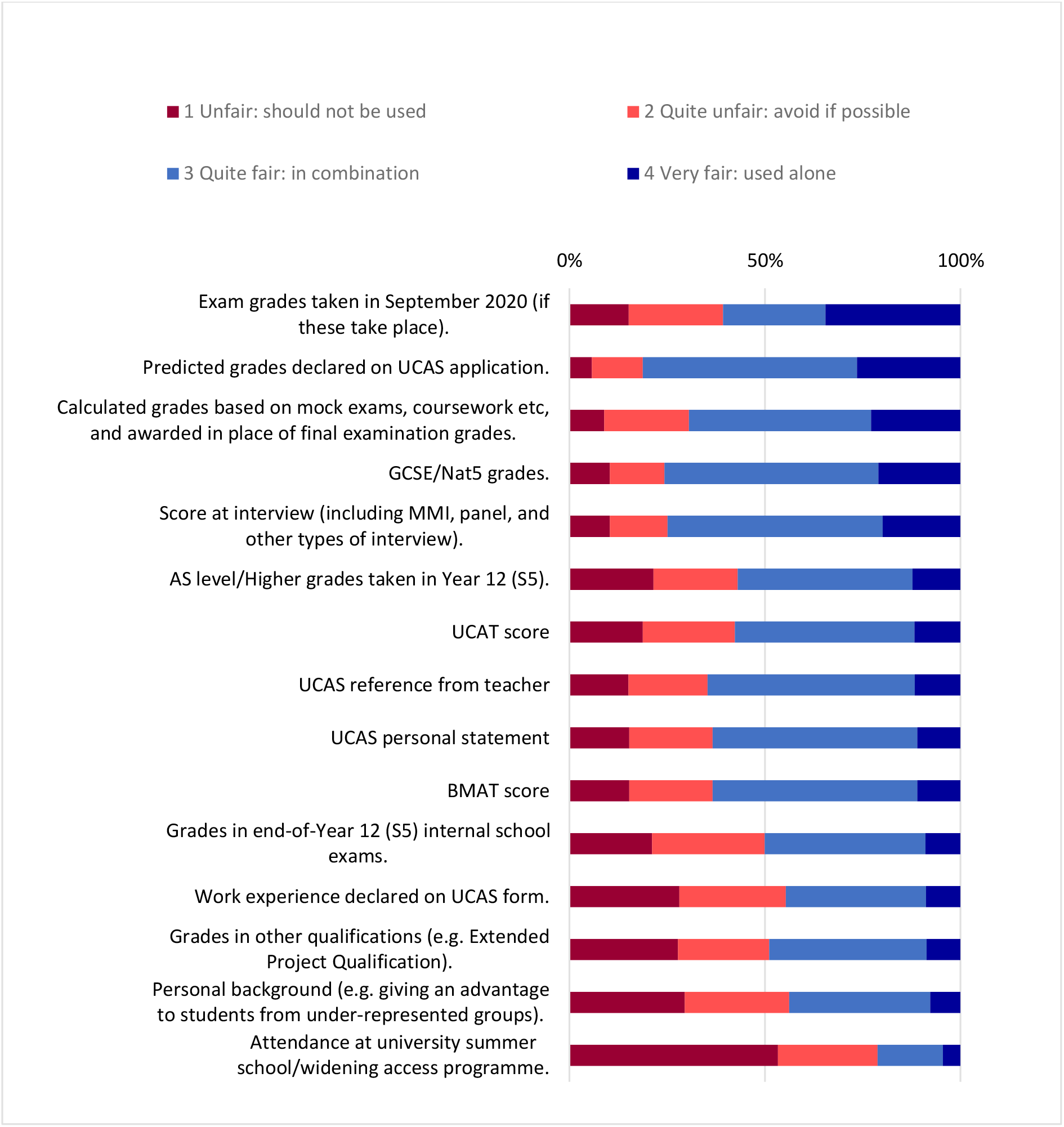
Perceptions of the fairness of methods medical schools could use to decide whether or not to accept applicants who currently hold an offer now that exams have been cancelled.

Multiple regression analyses showed that after taking account of all other educational and socio-demographic variables, BAME participants were more likely to perceive *Exams taken in September 2020, UCAS personal statement*, and *Personal background* as fair to use, and respondents from deprived areas were more likely to perceive *Personal background* and *Attendance at widening access programmes* as fair to use. *Calculated grades based on mock exams, coursework etc, and awarded in place of final examination grades* were perceived as less fair by those with lower predicted A-levels.

There were over 300 freetext responses, with participants elaborating on their responses or suggesting alternatives. Examples of elaborations include:

> “A combination of the most objective information that every offer holder will have, ie GCSEs, UCAT or BMAT, interview score, etc”
>
> “A standardised form of assessing all medical applicants would be the best way to allocate existing places. One could argue that most of us have already taken a standardised assessment, the UCAT. Since we do not have standardised A level grades, places should be offered using the UCAT as this is the fairest way of distributing places to the most able students.”
>
> “Using interview scores and UCAT scores in combination are independent measures, and are more fair than using calculated grades which have the potential to be biased.”
>
> “Anything including personal statement, BMAT or UCAT I would argue are unfair to use as judgement as there will definitely be a bias in terms of how certain students achieved their grade. I believe the fairest way to determine ones overall grade would be to use their GCSE data with a combination of evidence throughout the two years of A levels.”

Other measures participants mentioned included: an additional university assessment (written, viva or project/portfolio-based) now or at the start of the academic year, an additional interview, selection at the end of Year 1/make first year a foundation year, additional reference from teachers/school, reference from work experience, school/college attendance record, distance from university, extenuating circumstances, self-reported use of time during quarantine/lockdown, number of offers received, prioritise those with higher degrees, prioritise those already working in the NHS, extra-curricular achievement (e.g. music, Duke of Edinburgh’s Award), school’s prior achievement. For example:

> “NHS experience ie patient facing health professional ie years and grade, other non technical skills, education background ie. science, post graduate achievement ie MSc particularly if in science or medical subject and grade achieved. Also emphasis on the candidates as a whole ie well rounded personality (potential to communicate well) rather than typical A Grade student. Letter of recommendations from medical consultant whom candidates may have worked closely with.”
>
> “Another interview possibly over the phone to see what students have done with their time in quarantine (ie, volunteering in a care setting or hospital / working in a hospital/ exploring other interests)”
>
> “Each university could form their own selection test similar to UCAT/BMAT with a brief guidance/specification on what will be on the test given out to offer holders so they have some time to revise for it, but this should be used in combination with other details (e.g. if offer holder’s calculated grade was only 1 grade below what was required for entry)”
>
> “I think a combination of previous results, any exams that do go ahead (at some point whether that is this summer or later), alongside medical applications, relevant work experience (as per personal statement and any other forms detailing this) and the applicant interview. Also potentially the medical schools could generate online admissions tests for students with conditional offers to generate a clearer view of a students capability and ability to comprehend and withstand the pressures of medical school. But any tests generated by the medical schools must be used alongside the other parts of the applications to ensure fairness.”

Participants were asked whether they had heard anything from medical schools/universities they had applied to about how selection might be impacted by examination cancellations; among those holding conditional offers, a minority (n=538; 42%) said they had heard from at least one medical school/university they had applied to.

#### Acceptability of options for dealing with a situation in which more students meet their offers than there are medical school places

Participants were asked to rate the acceptability (“completely unacceptable”, “slightly unacceptable”, “neutral”, “slightly acceptable”, “completely acceptable”) of a number of options that medical schools could use if they had more students meeting offers than they had places.

The most acceptable option was *Ask some applicants with offers to volunteer to defer a year* which 38.5% participants felt would be completely acceptable and 28.8% felt would be slightly acceptable. The only other acceptable option was *Accept all applicants whose calculated grades meet the conditional offer, although it could mean fewer resources per student*, which 35.4% felt would be completely acceptable and 36.6% felt would be slightly acceptable. See **Figure 3**.

Multiple regression analyses showed no significant differences by social or demographic group on these items.

**Figure 3:**
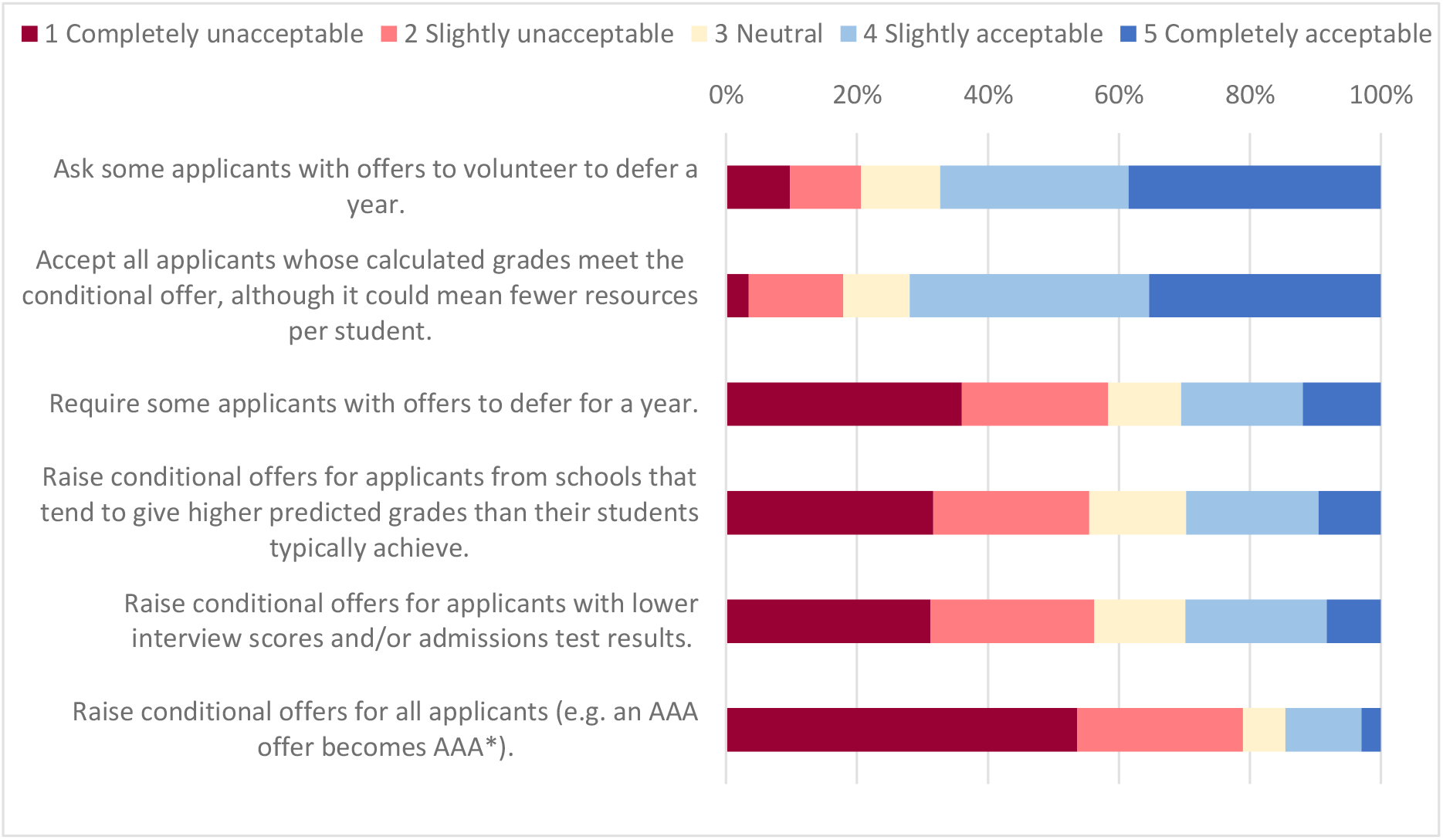
Acceptability of actions medical schools could take if they have more applicants meeting offers than they have places.

In freetext responses several respondents suggested that medical schools should receive more funding to manage larger cohorts and create more doctors, e.g.:

> “Deferring of one year should not be taken into consideration as this would damage applications of next year. Ask the government to invest more money on the NHS and allow to have more spaces. All these problems could be solved if exams were taken virtually.”
>
> “The government could also provide more funding for medical schools- not only will this allow more people to attend but it will also mean there are more doctors down the line who can work in the NHS.”

There were suggestions that applicants could opt to attend other medical schools they had applied to but which they had not selected as their firm or insurance choice, or that they could be offered places at medical schools they had not applied to:

> “If some medical schools have a lower numbers of applicants overall, compared to others, redistribute some students to these ones, with permission.”

There were many suggestions of incentives to defer, and some felt that they would welcome a year off before starting:

> “Incentives to defer like 1 yr free accommodation or £5000 or student ambassador job for gap year”
>
> “Incentive to deferring such as free university accommodation for the first year, organised work experience placements and or organised care assistant jobs for the gap year.”
>
> “If people are asked to volunteer to or forcefully defer entry, offering alternatives for work they could do within a healthcare setting for that year. For example, maybe clerical work within the NHS so they’re still immersed within the healthcare system.
>
> “Asking students to voluntarily defer a year would be a popular option, I think many people will reevaluate their priorities over the coming months and may appreciate the opportunity.”
>
> “The option to defer is definitely an option that should be considered as many people would be happy with the idea of gaining more medical experience in the year out that they would now have.”

There were suggestions medical schools could have multiple cohorts either all starting in October or one cohort starting in October and another cohort starting early 2021.

> “Create an extra group/year for Covid Students to manage the numbers”
>
> “Maybe consider having staggered starts throughout the year October start January start June starts.”
>
> “Stagger the course to offer two presentations and alter the following academic term holidays if possible”

Respondents also expressed concern as to the impact of the present disruption on next year’s admissions cycle and available resources:

> “The selection process should not be biased towards those rejected this year, next year, and should not change for the next cohort.”
>
> “I hope that this year’s or next year’s applicants will not be disadvantaged due to these unprecedented circumstances.”

#### Perceptions of potential impact on admissions for 2021

Participants were asked to rate how much they agreed or disagreed with six options as to how medical schools could deal with the potential impact of the current situation on admissions in 2021. See **Figure 4**.

**Figure 4:**
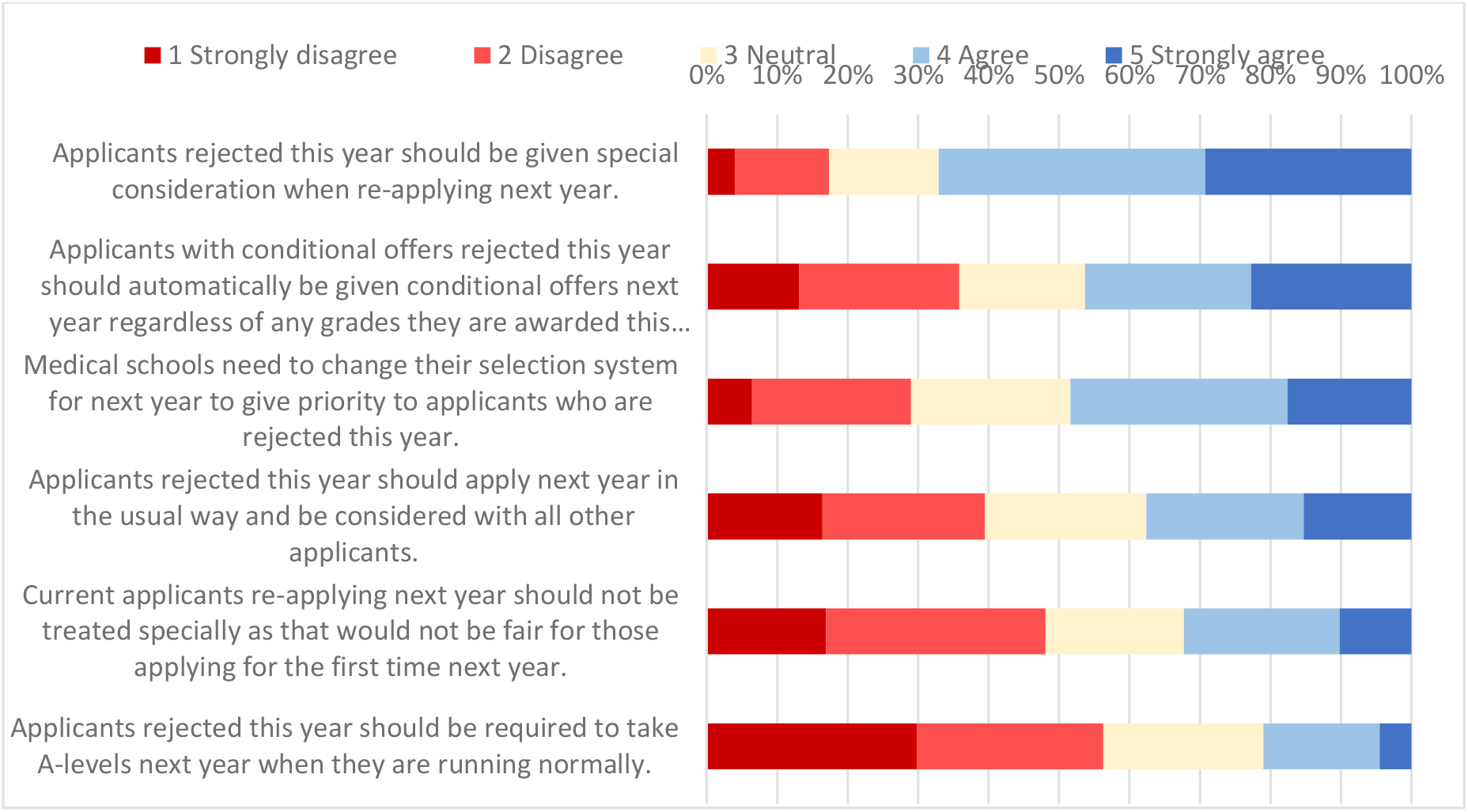
Views on how current applicants should be considered by medical schools if they reapply next year.

In general, respondents felt medical schools should give special consideration to current applicants reapplying next year (67.1% agreed/strongly agreed that *Applicants rejected this year should be given special consideration when re-applying next year*) however opinions were divided about what that special consideration should consist of.

Although 48.4% agreed/strongly agreed that *Medical schools need to change their selection system for next year to give priority to applicants who are rejected this year*, 29.0% disagreed/strongly disagreed. About a third of respondents agreed/strongly agreed that *Current applicants re-applying next year should not be treated specially as that would not be fair for those applying for the first time next year*, agreed/strongly agreed that *Applicants rejected this year should apply next year in the usual way and be considered with all other applicants*, and disagreed/strongly disagreed that *Applicants with conditional offers rejected this year should automatically be given conditional offers next year regardless of any grades they are awarded this year*.

Most participants (56.3%) did however disagree/strongly disagree that *Applicants rejected this year should be required to take A-levels next year when they are running normally*, with only 21.0% agreeing/strongly agreeing that they should be required to take them.

Multiple regression analyses showed that after accounting for number of offers, educational, social and demographic factors, BAME respondents were more likely to feel that re-applicants should be given some advantages.1 Strongly disagree

#### Starting academic year 2020/2021

A majority of respondents (n=952, 61.1%) believed that if necessary, medical schools should *Defer the start of the academic year only when face-to-face teaching is possible* with 605 respondents (38.9%) believing that medical schools should *Start the academic year on time using distance learning for as long as is necessary*. This did not vary significantly by prior attainment, number of offers, or educational/social/demographic background.

### Education and university preparation

#### Calculated grades and the perceptions of process of awarding calculated grades in lieu of examination grades

Participants were generally fairly ambivalent towards calculated grades. On the positive side (see **Figure 5 a**)), the majority of respondents (78.6%) preferred calculated grades to taking examinations next year, and about half (54.9%) preferred calculated grades to taking examinations in September 2020. Over half (59.3%) agreed that schools wouldn’t be able game the process to award all their students high grades, and 51.4% felt that the process of awarding calculated grades was the best way to be fair to most students in the circumstances (although 35.0% disagreed). Over half (56.4%) agreed that their teachers were generally able to rank and grade students accurately, however respondents were divided as to whether their own teachers knew them well enough to grade and rank them accurately: 42.0% agreed their teachers did NOT know them well enough whereas 44.6% thought their teachers DID know them well enough.

**Figure 5:**
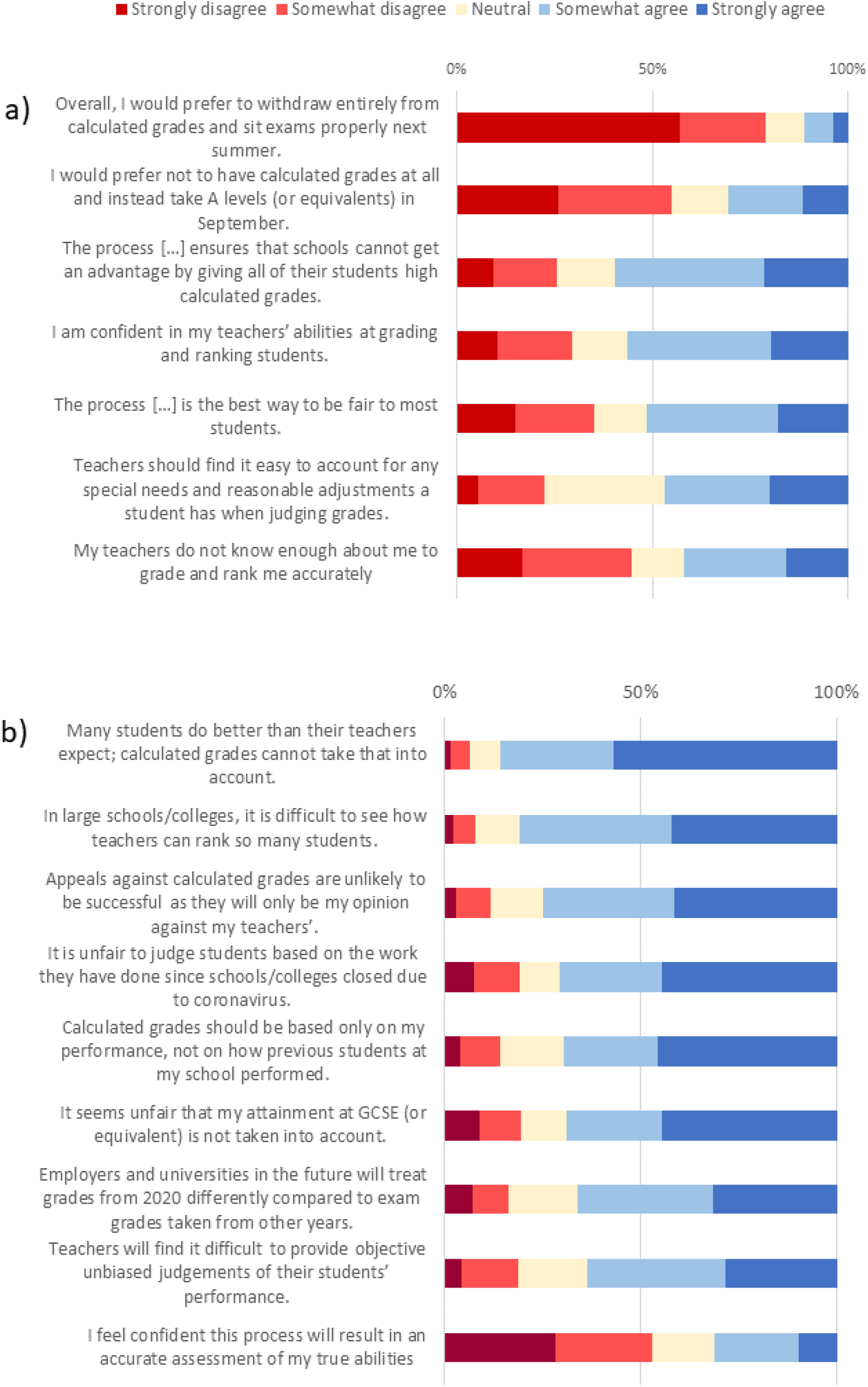
Aspects of calculated grades that respondents were generally more a) positive and b) negative about.

On the negative side (see **Figure 5 b**)), over half of respondents (52.9%) disagreed or strongly disagreed that calculated grades would result in an accurate assessment of their abilities, with 63.4% agreeing that teachers would find it hard to be unbiased, 80.7% agreeing it was difficult to see how teachers in large schools can rank so many students and 85.5% agreeing calculated grades cannot take into account students doing better in exams than their teachers expected. Most agreed it was unfair to judge students on work done since schools/colleges closed (70.4%), that grades should be based solely on their performance and not the performance of previous students at their school (69.6%), and that it was unfair their GCSE performance was not taken into account (68.7%).

Mean top three predicted A-level points was a major predictor of perceptions of calculated grades but there were also differences by background after accounting for prior attainment, number of offers and other educational/social/demographic factors: BAME respondents and female respondents were more negative about calculated grades and respondents from non-selective state schools and those from more deprived areas were more likely to agree that calculated grades should not take into account the performance of previous pupils at their school. See **Figure 6**.

**Figure 6:**
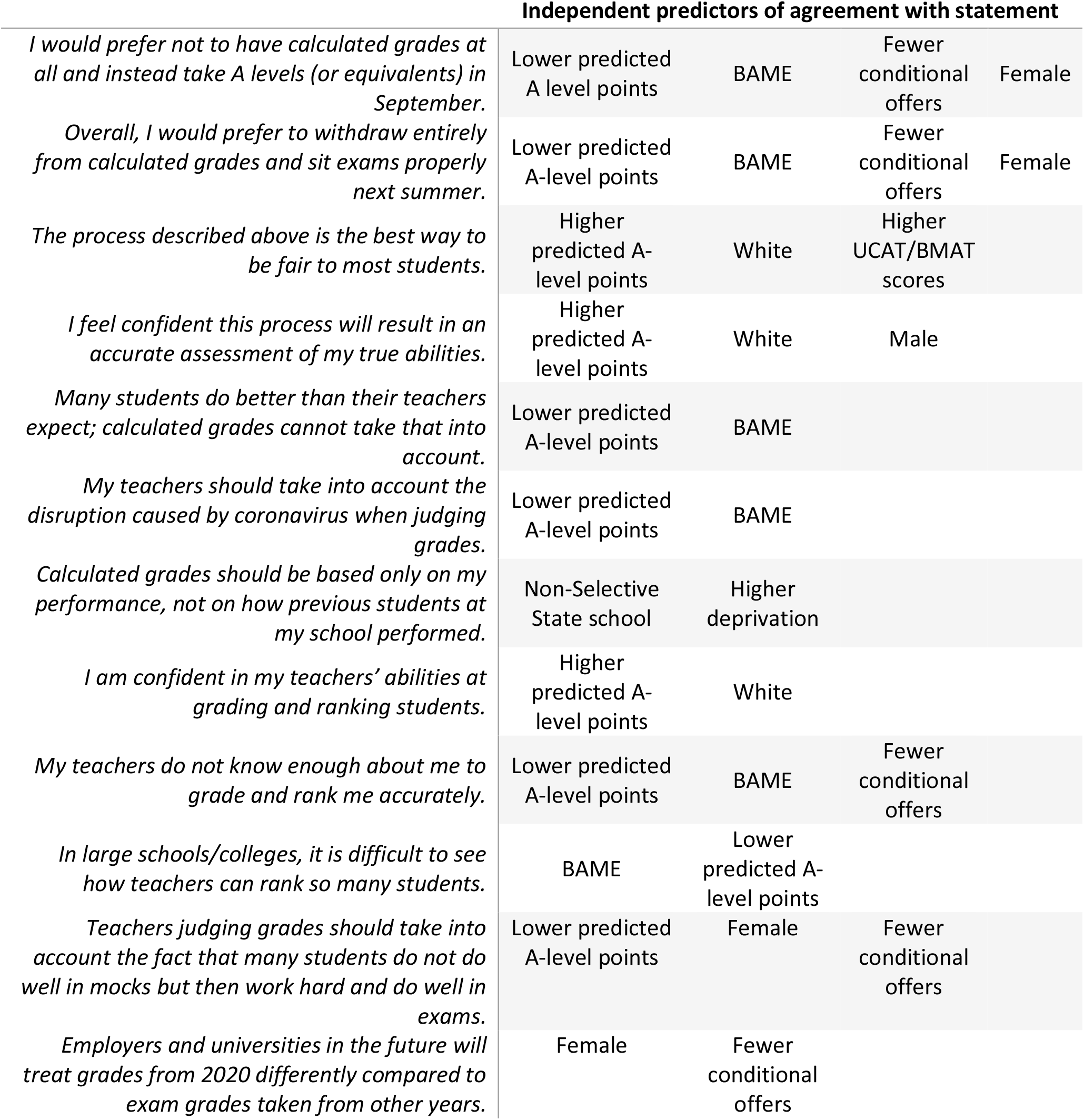
Predictors of agreement with statements relating to calculated grades. Predictors are ordered left to right by strength of relationship to the statement. Only statements that showed significant differences by social/demographic group after controlling for prior attainment and the number of offers are shown.

Freetext responses showed concerns that calculated grades would be based on work completed early in the academic year and on mock exams created and assessed by the school. It was felt that these measures would not take into consideration the development and academic progress made by pupils over the year, even when teachers gave special consideration to the impact of the disruption. There was also concern that at the time of mock exams in particular, many medicine applicants were more focused on admissions tests (BMAT in particular), submitting applications and preparing for interviews.

> “Grade calculations took away the chance the students had to prove themselves (final exams) and their control. Basing the final grade on a time when the students weren’t aware that they were being truly assessed can hardly be classed as fair.”
>
> “I believe universities should be lenient and realise that if a students calculated grade is below their conditional offer, this is not 100% representative of the students abilities. If they were able to secure an offer in the first place then universities should already know the academic capabilities of said student through their GCSE grades, predicted grades, UCAT/BMAT scores, teacher references, interviews etc. Otherwise, they wouldn’t have given the student an offer. Where possible, every offer holder should be given their place at university in this academic year, whenever it resumes and should not be forced to take a year out and spend that year being stressed, lost and demotivated.”

With teacher submitted grades then being subject to standardisation by the exam boards based on previous achievement from a school was a concern for this student:

> “I am the only student in my year and the third student in my sixth form’s history to ever apply for medicine, and the first to receive all 5 offers. My school historically is one that does not do very well and I fear that my individual success and all the hard work I have had to do on my own as I get no help from my school, will be overshadowed by the bad results from previous years.”

#### Education since the shutdown

A minority of respondents said their school was planning on formally assessing them on work done since the shutdown (n=184; 11.8%); nearly half (n=740; 47.5%) said their school would not, and over a third (n=614; 39.4%) were uncertain. Respondents attending a private/selective school were twice as likely to report being assessed on work since the shutdown (14.2% vs 7.6%; p<.001).

Participants were asked whether they were using educational resources provided by their school/college and if not why not. Nearly all respondents had used at least one resource (n=1346; 91%) and three was the average number used.

Respondents attending private/selective schools were more likely to report having used all educational resources except support for university applications, and those at non-selective state schools used on average two resources compared to the three used by those at private/selective schools. The largest difference was in the use of online teaching in real time, which those at private/selective schools were nearly four times more likely to have used. See **Figure 7**.

**Figure 7:**
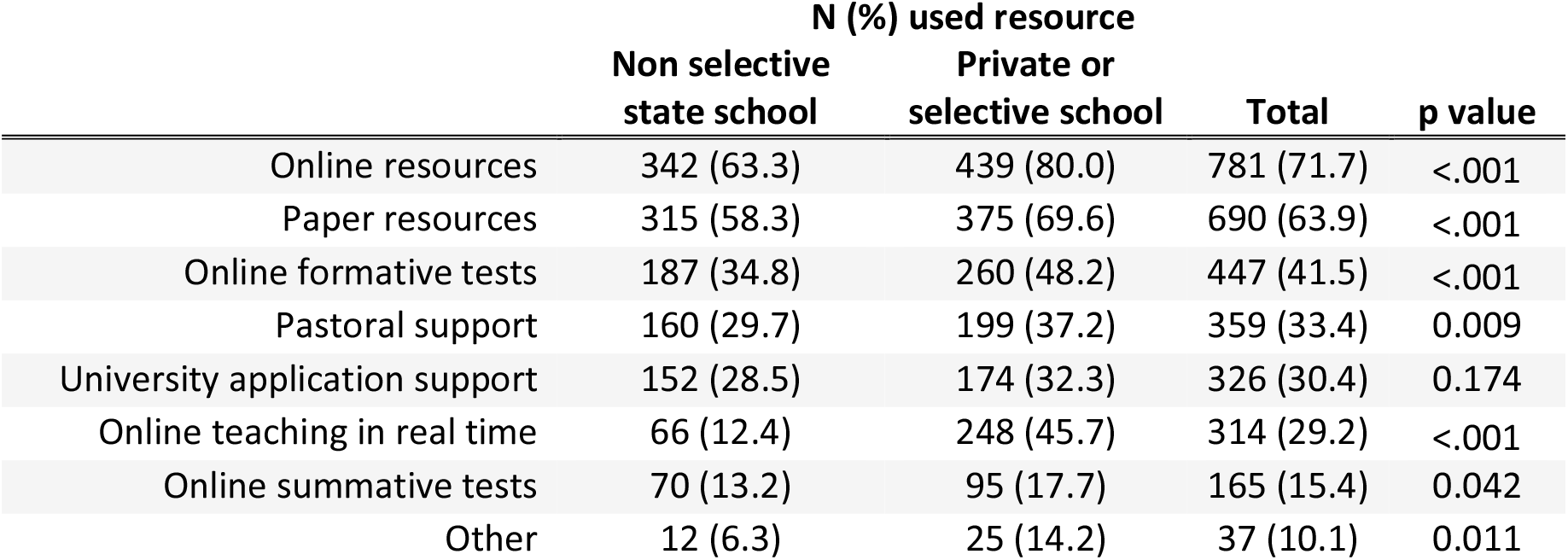
School-provided educational resources used by respondents from non-selective state schools and private/selective schools.

In the multivariate analyses, attendance at a private/selective school was an independent predictor of using online teaching in real time, online resources for home learning, online formative assessments, and paper resources for home learning, even after controlling for prior attainment and socio-demographics. In addition, having at least one parent/carer with a university degree was an independent predictor of using paper resources for home learning, and having lower UCAT/BMAT scores was an independent predictor of using online teaching in real time.

Those who had not used educational resources reported the main reason(s) were either that the resources were not available or that they felt they did not need to use them. Only very few said they had not used a resource because of a lack of private quiet space, lack of time, lack of internet/computer access, or because they were finding it too hard to focus. Those at non-selective state schools were more likely than those at private/selective schools to state lack of availability as a reason, and less likely to state not needing to as a reason– see **Figure 8**.

**Figure 8:**
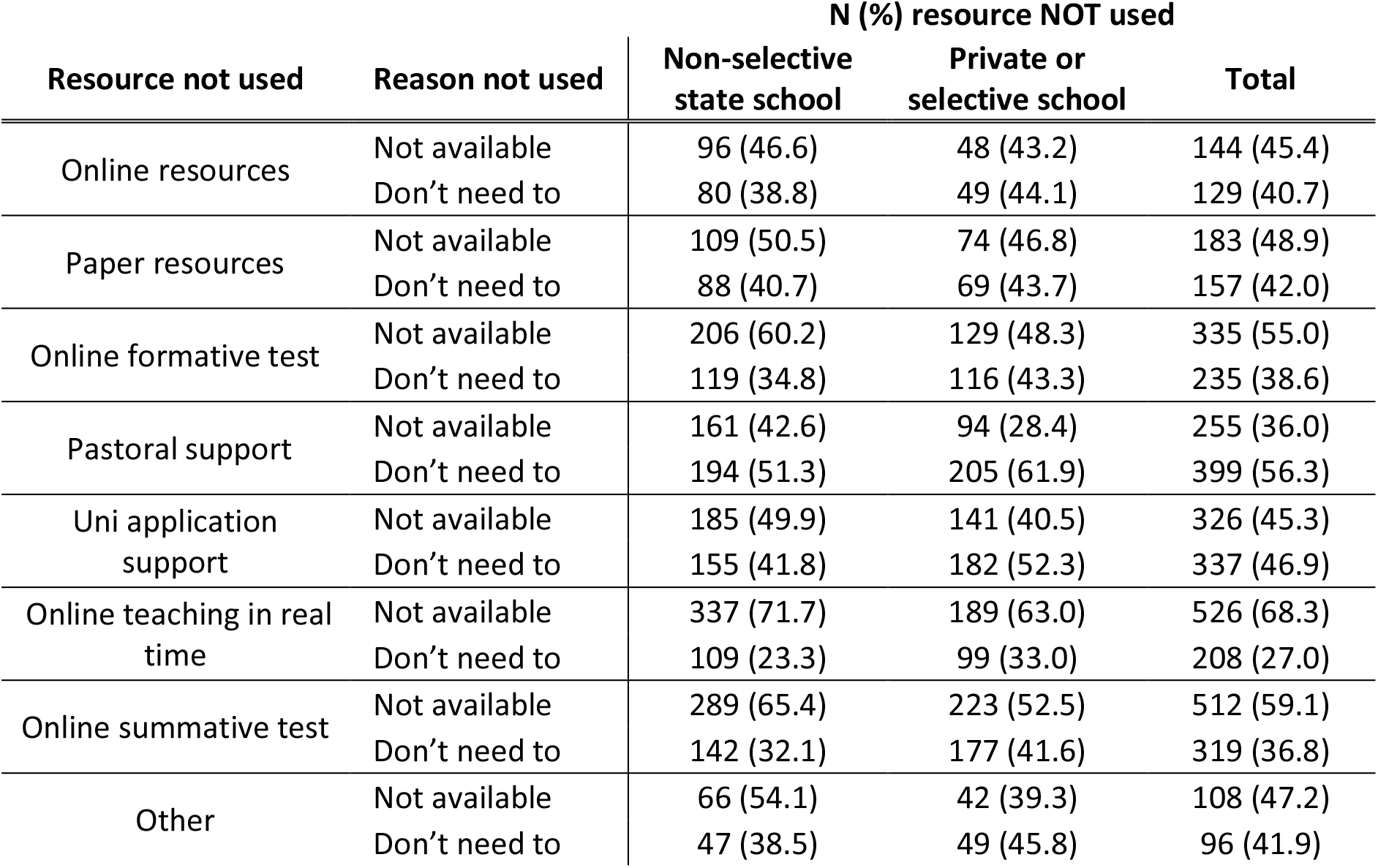
Respondents’ main reasons for not using school educational resources during the shutdown by school type.

#### Preparation for medical school/university

Participants were asked what preparation if any they were doing for university or medical school – see **Figure 9**.

**Figure 9:**
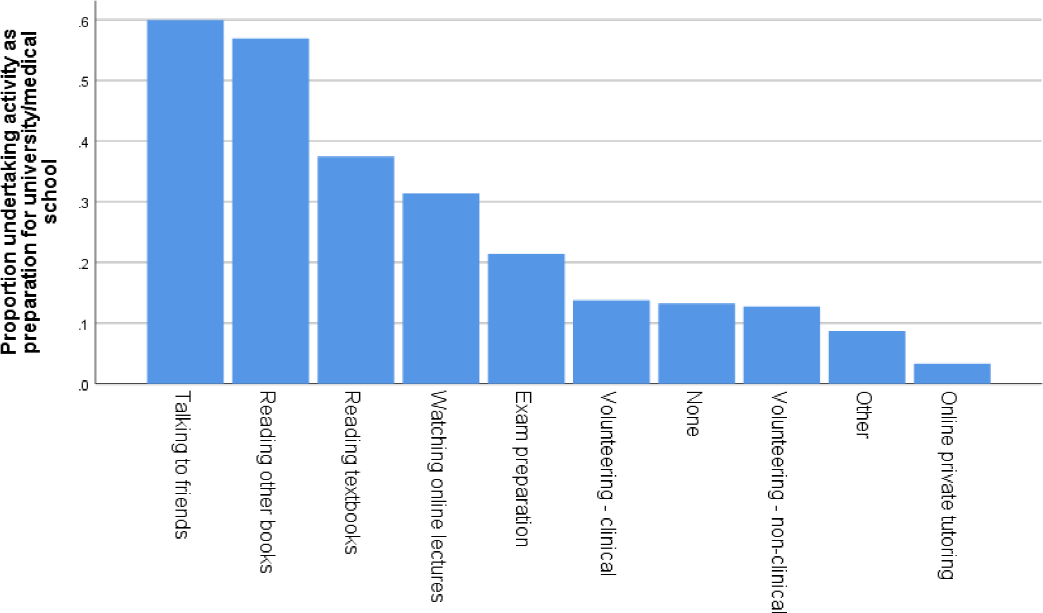
Proportion of respondents undertaking various activities to prepare for medical school or university.

Of the 207 (13.3% of the sample) who said they were not doing any preparation, the most common reason was that they were too worried and couldn’t focus (n=88; 42.5% of those not doing any preparation), not having resources (35.5%), feeling it wasn’t necessary (29.5%), caring for others (13.5%), not going to university this year (14.0%), not having time (6.3%), and being unwell (4.8%). Respondents could select multiple reasons.

### Time spent during the lockdown

Participants were asked to state how much time they were spending on various activities in the previous five days – see **Figure 10**. The multivariate analysis showed that respondents from private/selective schools reported spending more time studying, even after controlling for prior attainment and socio-demographic factors.

**Figure 10:**
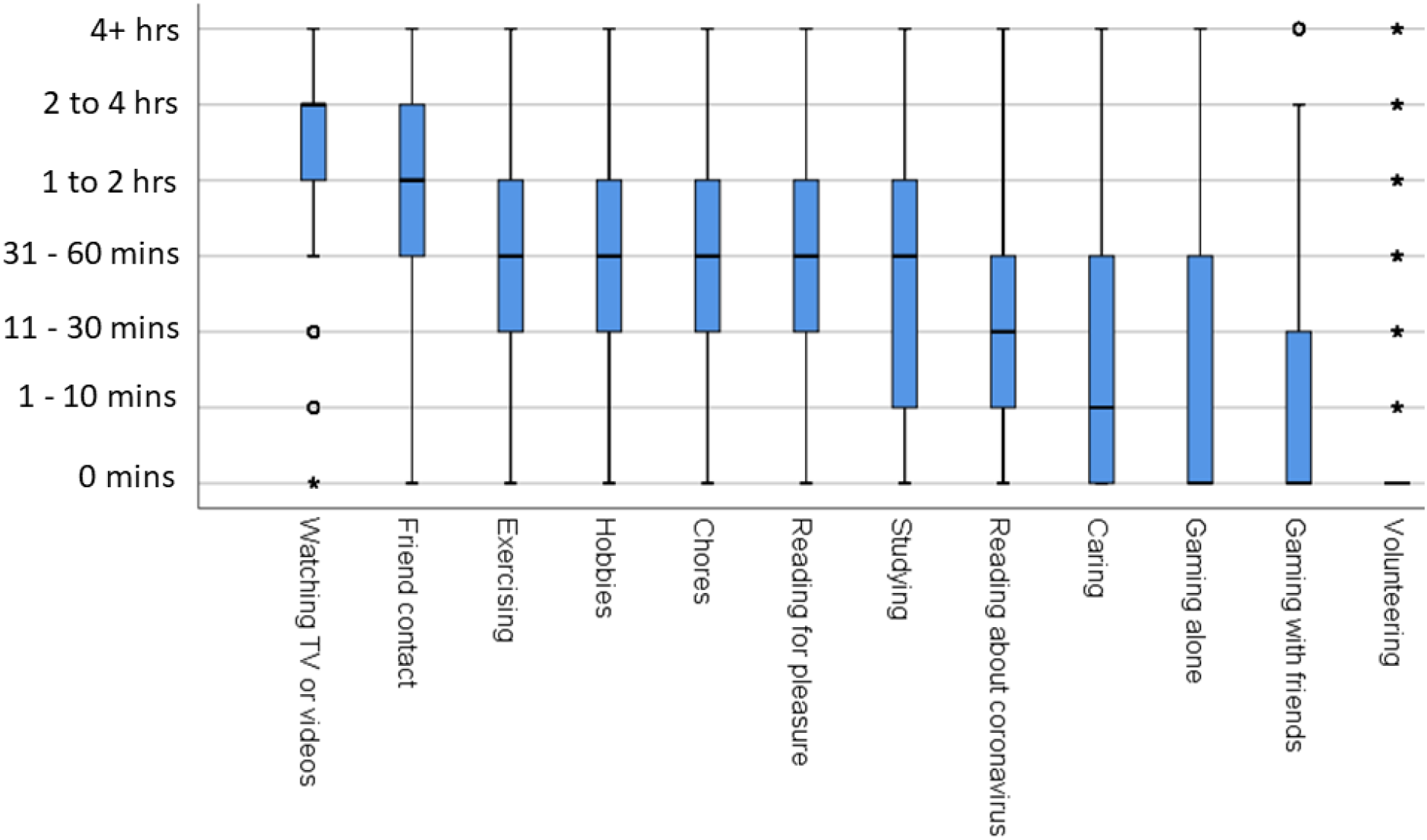
Amount of time respondents reported spending on various activities during the lockdown.

#### Personality measures and time spent during lockdown

Personality traits are “relatively enduring styles of thinking, feeling, and acting”.(19) It is generally agreed that there are five distinct personal traits or factors: Neuroticism, Extraversion, Openness to Experience, Agreeableness, and Conscientiousness. Correlations between personality and time spent on activities are shown in **Figure 11**.

**Figure 11:**
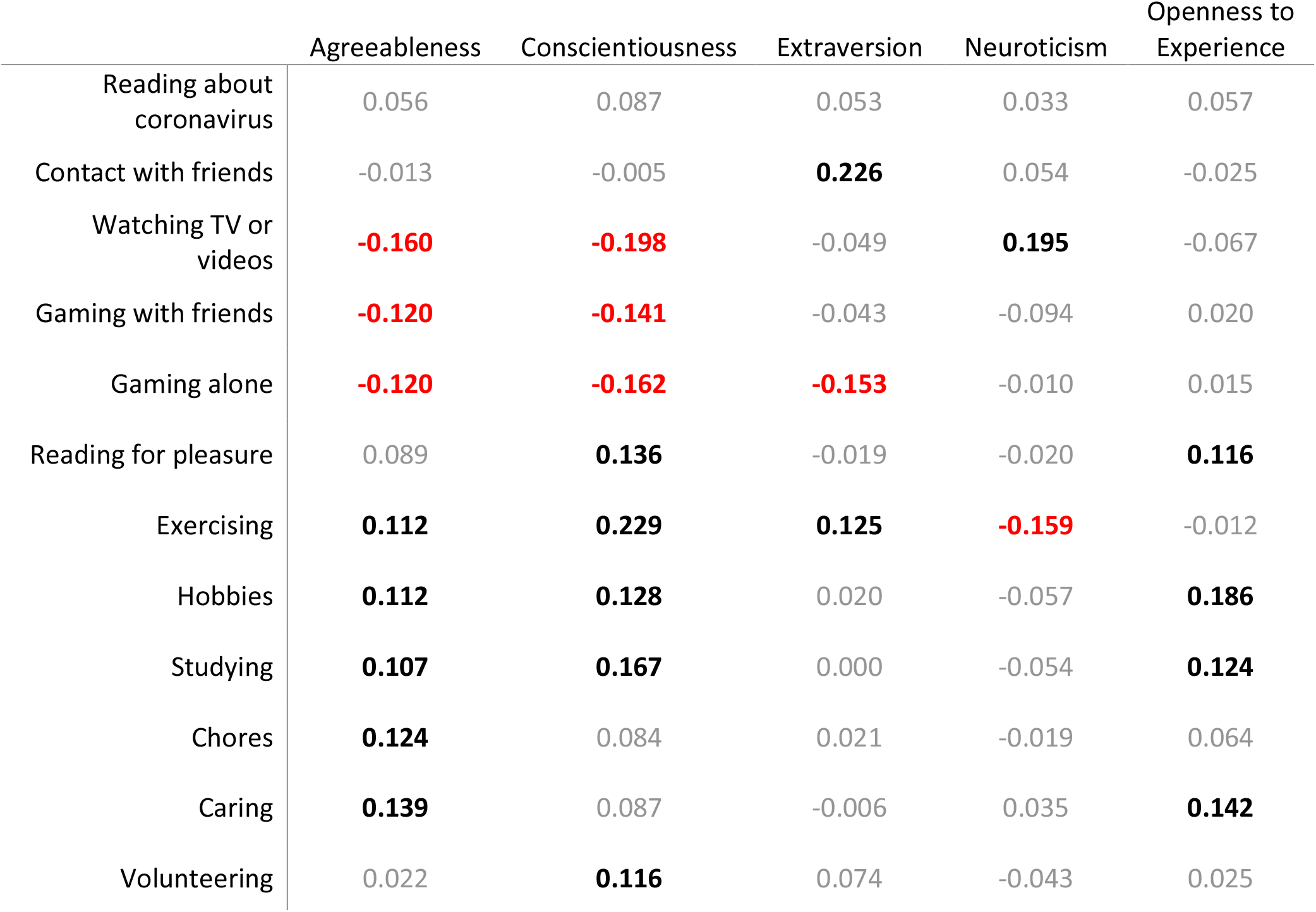
Simple correlations (Pearson’s r) between time spent on various activities during lockdown and big five personality traits. Correlations in bold are significant at p<.001: those in red are negative and those in black are positive.

## Summary and conclusions

### Summary of results

No single measure, including calculated grades, was considered fair enough by most applicants to use in the acceptance or rejection of offer-holders; however many applicants considered calculated grades – and many other measures - fair enough to use in combination with other measures such as interview scores or admission test scores. Taking into account personal background or widening participation attendance was considered fairer by BAME applicants, those from deprived areas, and those without degree-educated parents.

Many respondents had concerns about calculated grades, especially BAME and female applicants who felt teachers would find it difficult to grade and rank students accurately, and those from non-selective state schools and living in deprived areas were more concerned about the standardisation process that uses the attainment of previous pupils at a school. Despite this, the majority would rather have calculated grades than forgo calculated grades completely and take examinations in Autumn 2020 or Summer 2021 instead.

Respondents mostly felt that medical schools should admit any applicant who met their conditional offer, even if that meant having to increase the number of places (which would require a legal change and increased government funding), although there was also acceptance of medical schools asking for volunteers to defer but not of requiring deferrals. Respondents were divided as to how rejected applicants should be treated if they were to reapply next year, with some respondents feeling they should be treated no differently and others feeling their 2020 experience should be taken into account. A majority of respondents tended to favour medical schools delaying the start of term until face-to-face teaching were possible.

Applicants from non-selective state schools reported using fewer educational resources than their counterparts at private or selective schools, and in particular they reporting less online teaching in real time, and spending less time studying during the lockdown.

### Comparisons with other research

Our findings show many similarities to other recent UK studies of the effects of the COVID-19 pandemic on education in the UK (8, 9, 15, 16) however it is notable that in this sample of medical applicants ethnicity is more significant than socioeconomic factors in predicting concerns about calculated grades – indeed, to our knowledge, ours is the only survey of applicants that includes a measure of ethnicity. It is known that predicted grades are lower for some minority ethnic groups (20) and indeed, on 2^nd^ April 2020 after the announcement of the cancellation of examinations but before *Ofqual* specified details of calculated grades, the Runnymede Trust and several other race equality organisations wrote to the Secretary of State for Education to urge him to “ensure a fair, transparent and robust system which will more accurately reflect the ability and attainment of students from different backgrounds”.(21) Subsequently, on 30^th^ April, the Equality and Human Rights Commission said that,

> “Using predicted grades in place of this year’s summer assessments could deepen the existing inequality in education and put the future of disadvantaged young people at risk if not correctly implemented” (22)

Our finding that students from private/selective schools were using more educational measures - especially online teaching in real time, which requires significant teacher input and which Andrew et al (15) argue is higher quality that other types of resource - reflects findings from those authors’ research with parents of secondary school children (15) and teachers (16); however in our sample students’ use of educational resources and time spent studying did not vary by socioeconomic background, including parental higher education, socioeconomic status, or area deprivation. This may be a feature of this particularly high-achieving sample of medical applicants.

### Strengths and limitations

This study is, to our knowledge, the first systematic exploration of medical applicant views on and experiences of the most significant changes to UK education in living memory. It is also the first study we are aware of that looked at university applicant views on The large sample size gathered from around the UK, and the richness of the data allowed us to examine important differences in the experiences and views of different socio-demographic groups, after controlling statistically for educational attainment.

The speed at which we were required to develop the questionnaire and the unprecedented nature of the topic under investigation meant we were unable to use validated measures for most questions, nor have we been able to validate the measures ourselves, although we were able to pilot them with current applicants.

It is uncertain how representative our sample is of all medical applicants. Data on applications, offers, acceptances and academic achievement from the current UCAS cycle are not released until early 2021, but it is very likely that offer-holders were over-represented in our sample. Data from the 2019 UCAT testing cycle also show that our sample scored higher than the mean [https://www.ucat.ac.uk/media/1329/2019-test-statistics-oct-2019.pdf]; however not all UCAT test- takers apply to medicine. Demographic data on 2020 medical applicants released by UCAS in November 2019 showed that our restricted sample was similar to all English applicants aged 17 to 19 in terms of ethnicity and deprivation but had more women [https://www.ucas.com/data-and-analysis/undergraduate-statistics-and-reports/ucas-undergraduate-releases/applicant-releases-2020/2020-cycle-applicant-figures-15-october-deadline].

Medical applicants are not representative of all university applicants in either academic or socio-demographic terms; however the similarity of some of our findings to that of other research, for example that private school pupils are receiving significantly more education than non-selective state school pupils, suggests that the views and experiences of our sample may not be completely different from those of university applicants more generally; however generalisations from our findings to all applicants should only be done with extreme caution.

### Implications for policy and practice

The impact of calculated grades on medical admissions is uncertain. Our questionnaire closed on 22^nd^ April and on 5^th^ May 2020 the Medical Schools Council announced that medical schools would honour all offers met - meaning this was not clear at the time of our questionnaire - while acknowledging that there were still a number of issues that needed resolving.

How calculated grades are likely to work in practice has also been explored by a parallel analysis by our team using UKMED data over the last ten years, comparing predicted A-level grades (which are likely to be similar to calculated grades) with actual, attained A-level grades.(23) Predicted grades were systematically higher in medical school applicants than eventual achieved grades. In addition the predictive validity of predicted grades was only about two-thirds that of achieved A-level grades, both for outcomes five or six years later at the end of medical school, and seven or eight years later in postgraduate examinations. The under-prediction by predicted grades was mitigated in part, although not entirely, by combining predicted grades with UCAT/BMAT scores, which supports the views of some applicants that other measures might be used for selection amongst applicants not meeting the terms of conditional offers.

The likely impacts on medical schools of using calculated grades are uncertain, but our estimates suggest that there could in effect be a lowering of entry grade requirements, with possible subsequent increases in medical school drop-out rates, and a somewhat academically weaker cohort with poorer performance in medical school and postgraduate examinations.(5, 24) That is potentially important since very poor postgraduate examination performance itself strongly predicts being sanctioned by the medical regulator.(25)

In the awarding of calculated grades, the raw ‘centre assessment grades’ and rankings produced by teachers for *Ofqual* are likely to be similar to predicted grades in being more generous than achieved A-level grades would have been, although the standardisation to be used by examination boards and *Ofqual* are likely to minimise that effect, so that distributions of calculated grades within subjects and centres become similar to actual A-level grades in previous years. There are however still many uncertainties concerning calculated grades, and it is unclear whether there may be either an excess or a deficit of candidates meeting conditional offers, and there may well be differences also in candidates applying via clearing and related processes.

In this questionnaire many applicants felt it could be fair to using other information such as interview score, UCAT score, or GCSE score to accept or reject offer-holders, and this could include in selecting from amongst ‘near-misses’. Overall respondents to our questionnaire demonstrate a lack of confidence in the process of calculated grades. Given the concerns of the Equality and Humans Rights Commission, and the clear concerns also expressed in our study by some disadvantaged groups, there is a clear need to ensure that entrants as far as possible continue to reflect the breadth of those applying to study medicine.

Were there to be an excess of candidates meeting conditional offers as a result of calculated grades then applicants in our study suggested that that, in light of the shortage of doctors,(26) medical schools might argue for increased places and funding, although that might not be straightforward, either in financial terms, or in predicting through to numbers of places for clinical teaching, foundation training and so on. It is also worth considering that cohort sizes at many medical schools are already very large, that students tend to be less satisfied at larger schools,(27) and that accommodating extra students into face-to-face teaching that is COVID-secure is likely to be extremely challenging. These downsides would need to be balanced against the need for more doctors and the benefits of what would likely be a more socially and demographically diverse cohort.

The cancellation of public examinations and the use of calculated grades are not the only problems facing the 2020 application cohort. They are also at risk, particularly those from non-selective state schools, of coming to medical school having had less education over the previous few months,(14) meaning medical schools may need to provide additional teaching and resources to help students catch up. This is likely to be especially challenging for medical schools given the huge constraints on university budgets arising from drops in student numbers(28) and given that many are likely to be unable to open for face-to-face teaching at the start of the academic year, which in itself has unknown consequences.

The 2020 cohort of entrants is likely to face more uncertainty than any cohort of medical student entrants in the past half-century, and our survey makes very visible the many concerns of those applicants.

## Conclusions

The global tragedy of the coronavirus pandemic, in addition to its extensive mortality and morbidity, has resulted in huge and sudden disruptions to established ways of life including education and training at all levels. Medical education and training is no exception. The coronavirus pandemic will have significant and long term impacts on the selection, education and performance of our future medical workforce. Understanding how medical education will be affected is therefore important, and in particular how applicants to become the newest entrants to medical careers are being affected. Now more than ever we need medical education, and medical education research, to be prioritised and funded so we can ensure our future doctors are able to be resilient, successful and happy healthcare professionals providing excellent patient care. The present study provides a wide range of insights into the feelings of the 2020 cohort of applicants, only a small proportion of which we have adequately been able to report here.

## Data Availability

The data will be linked into the UK Medical Education Database www.ukmed.ac.uk to which researchers can apply for access.

## Acknowledgements

Firstly we are immensely grateful to the several thousand medical school applicants who took the time to respond to survey with a very tight time window, and we particularly thank those who commented that they were pleased that the survey they gave them an opportunity to express their thoughts, feelings and anxieties. We could not include everything that was said, but all comments have been read by the team, and we hope that the current paper summarises some of those many and varied views. We are also grateful to Paul Garrud, Clare Owen, Konstantinos Lulo, and Ewan McNichol for their comments on earlier versions of the questionnaire.

## Contributors

KW, DH and ICM jointly developed the idea for the study, and developed the questionnaire together. DH was responsible for putting the questionnaire online, and for identifying applicants to whom it should be sent, as well as sending text and email reminders. DH and KW cleaned the data, and KW, DH and ICM were all involved in data analysis. The report was written jointly by all three authors, and all authors have read and reviewed the final draft.

## Disclaimers

KW and DH state that this publication presents independent research funded by the National Institute for Health Research (NIHR). The views expressed are those of the authors and not necessarily those of the NHS, the NIHR or the Department of Health and Social Care.

## Provenance and peer review

Not commissioned; to be submitted to an externally peer reviewed journal.

## Data sharing statement

The data will be linked into the UK Medical Education Database **www.ukmed.ac.uk** to which researchers can apply for access.

## Footnotes

1 See **Figure 2** for full item wording.

